# Common genetic variations in telomere length genes and lung cancer

**DOI:** 10.1101/2022.08.24.22279131

**Authors:** Ricardo Cortez Cardoso Penha, Karl Smith-Byrne, Joshua R Atkins, Philip Haycock, Siddhartha Kar, Veryan Codd, Nilesh J Samani, Christopher P Nelson, Maja Milojevic, Aurélie AG Gabriel, Christopher Amos, Paul Brennan, Rayjean J Hung, Linda Kachuri, James D McKay

## Abstract

**Background:** Genome-wide association studies (GWAS) have identified genetic susceptibility variants for both leukocyte telomere length (LTL) and lung cancer susceptibility. Recently, 108 novel genetic loci within genes involved in telomere biology and DNA repair have been linked to LTL in UK Biobank. In the current work, we investigated the relationship between genetically predicted LTL and lung cancer.

**Methods:** To explore the shared genetic basis between LTL and lung cancer, we performed genetic correlation, Mendelian Randomization (MR), and colocalisation analyses using the largest available GWASs of LTL (N=464,716) and lung cancer (29,239 cases; 56,450 controls). To further characterize the molecular mechanisms underlying this relationship, principal component analysis (PCA) was used to summarize gene expression profiles in lung adenocarcinoma tumours from The Cancer Genome Atlas.

**Results:** Although there was no genome-wide genetic correlation between LTL and lung cancer risk (r_g_=-0.01, p=0.88), MR analyses using 144 instruments identified a putatively causal association. Longer LTL conferred an increased risk of lung cancer (OR=1.62, 95%CI=1.44-1.83, p=9.9×10^−15^), lung cancer in never smokers (OR=2.02, 95%CI=1.45-2.83, p=3.78×10^−05^), and lung adenocarcinoma (OR=2.43, 95%CI=2.02-2.92, p=3.8×10^−21^). Of these 144 LTL genetic instruments, 12 showed evidence of colocalisation with lung adenocarcinoma risk and revealed novel susceptibility loci, including *MPHOSPH6* (rs2303262), *PRPF6* (rs80150989), and *POLI* (rs2276182). A polygenic risk score for LTL was associated with the second principal component (PC2) of gene expression (Beta=0.17, p=1.0×10^−3^). The aspect of PC2 associated with longer LTL was also associated with being female (p=0.005), never smokers (p=0.04), and earlier tumour stage (p=0.002). PC2 was strongly associated with cell proliferation score (p=3.6×10^−30^) and genomic features related to genome stability, including copy number changes (p=1.6×10^−5^) and telomerase activity (p=1.3×10^−5^) in the multivariate regression analyses.

**Conclusions:** This study identified an association between longer genetically predicted LTL and lung cancer and sheds light on the potential molecular mechanisms related to LTL in lung adenocarcinomas.

## Background

Telomeres are a complex of repetitive TTAGGG sequences and nucleoproteins located at the end of chromosomes and have an essential role in sustaining cell proliferation and preserving genome integrity [1]. Telomere length progressively shortens with age in proliferative somatic cells due to incomplete telomeric regions replication [2] and low activity of the telomerase *TERT* in adult cells. The shortening of the telomere length results in cell cycle arrest, cellular senescence, and apoptosis in somatic cells [3]. The maintenance of telomere length, which allows cancer cells to escape the telomere-mediated apoptosis pathway, is one of the hallmarks of cancer [4].

Telomere length appears to vary between individuals and has been studied in relation to many diseases. In observational studies, telomere length is measured as the average length of telomeric sequences in a given tissue [5]. Telomere length appears correlated across tissue types [6], and as such, leukocyte telomere length (LTL) is generally measured in epidemiologic studies as a proxy for telomere length in other tissues. Recently, LTL has been measured in 472,174 individuals from the UK Biobank (UKBB) [7] and LTL was associated with multiple biomedical traits (i.e, pulmonary and cardiovascular diseases, hematological traits, lymphomas, kidney cancer, and other cancer types). Genetic analysis of LTL also revealed 138 genetic loci linked to LTL across a variety of different genes involved in telomere biology and DNA repair [7].

In the context of lung cancer, genetic variants at several loci have been associated with both LTL and lung cancer risk, including variants near the *TERT, TERC, OBFC1*, and *RTEL1* genes, fundamental to telomere length maintenance [8,9,10,11,12]. The effects of the telomere-related variants appear more relevant to lung adenocarcinoma risk than other histologic subtypes [12,13]. Accordingly, a causal relationship between LTL and susceptibility to lung cancer was observed using Mendelian randomization (MR) approaches [14,15,16] as well as in observational studies that have associated directly measured telomere length with risk of lung cancer [17,18].

The aim of the current work was to investigate the relationship between genetically predicted LTL and lung cancer, including lung cancer histological subtypes and smoking status. To this end, we conducted genome-wide correlations, MR, and colocalisation analyses to explore the relationship between LTL and lung cancer. We additionally undertook polygenic risk score analysis using the LTL genetic instrument to explore the influence of LTL on the demographic, clinical, and molecular features of lung adenocarcinoma tumors.

## Material and methods

### Data

GWAS summary statistics for lung cancer (29,239 cases and 56,450 controls) and stratified by histological subtype (squamous cell carcinoma, small cell carcinoma, and adenocarcinoma) and smoking status (ever and never smokers) were obtained from the International Lung Cancer Consortium (ILCCO) [12]. All analyses of LTL requiring summary statistics used results from a GWAS of LTL in 464,716 individuals of European ancestry from the UKBB [7]. Downstream analyses considered additional lung cancer risk factors, such as lung function and cigarette smoking. We obtained GWAS summary statistics for forced expiratory volume in 1 second (FEV_1_) and forced vital capacity (FVC) from a published UKBB analysis [19]. For smoking behavior traits, we used results from the GSCAN consortium meta-analysis of cigarettes per day (continuous), smoking initiation (ever vs never), smoking cessation (successfully quit versus continuing), and age at smoking initiation (continuous) [20] excluding the UKBB participants. Colocalisation analyses of gene expression used lung tissue eQTL summary statistics from GTEx data version 8.

Analyses of molecular phenotypes were performed using 343 lung adenocarcinoma samples of European ancestry from The Cancer Genome Atlas (TGCA) cohort with both germline and RNA-sequencing data available. Genotyping and imputation of germline variants have been described elsewhere [21]. The total somatic mutation burden of TCGA samples were obtained from Ellrott et al., 2018 [22] and DNA mutational signatures were extracted and attributed, as previously described [21]. RNA-sequencing data were obtained from TCGA data portal using TCGAbiolinks package in R (version 2.22.3) [23]. Telomere length measurement by whole-genome sequencing (WGS-measured TL, 655 samples across cancer sites) was retrieved from Barthel et al., (2017) [24].

Tumor genomic characteristics were defined by the analyses of the TCGA data, including gene expression-based scores of telomerase activity [24] and cellular proliferation [25], as well as the observed frequency of somatic homologous recombination related events (represented as a homologous recombination repair deficiency score), and the average number of somatic copy number alteration within the tumors [26].

### Linkage disequilibrium score regression

Genetic correlations across traits were calculated using Linkage disequilibrium score regression (LDSC) by the LDSC package (v1.0.0) [27]. LD scores were generated on the 1000 Genomes Project Phase 3 reference panel with the HLA region excluded as provided by the package due to long range LD patterns. The genome-wide correlations that passed Bonferroni correction (adjusted p-values <0.05) were considered statically significant.

### Mendelian Randomization

MR is a method for interrogating relationships between putative risk factors and health outcomes by using genetic variants associated with the exposure of interest, typically obtained from GWAS, as instrumental variables. Assuming that fundamental MR assumptions are satisfied, this approach can be said to identify unbiased causal estimates. The genetic instrument for LTL was defined as the set of 144 genetic variants that were genome-wide significant (p<5e^-08^) but not in linkage disequilibrium with each other (r^2^ < 0.01) and restricted to common genetic variation (minor allele frequency > 1%). Proxy variants in LD (r^2^ > 0.8) were chosen when a genetic variant was not available in the lung cancer GWAS. Primary MR analyses were conducted using the inverse-variance method with multiplicative random-effects [28]. Sensitivity analyses to horizontal pleiotropy and other violations of MR assumptions were performed using other MR estimation methods, such as weighted median, MR-Egger, contamination mixture model, MR-PRESSO, and MR-RAPS [28,29]. Multivariable MR (MVMR) methods included the inverse-variance weighted, MR-Egger, and LASSO-based methods [28].

### Colocalisation methods

Unlike MR, where the goal is to assess the evidence for a causal effect of an exposure on an outcome, colocalisation is agnostic with respect to direction of effect and only assesses the probability that the two traits are affected by the same genetic variants at a given locus. Colocalisation can be viewed as a complementary approach for evaluating MR assumptions within specific genes or regions, since strong evidence of colocalisation indicates overlap in genetic mechanisms affecting LTL and lung cancer. We used COLOC (v5.1.0) [30] to estimate the posterior probability for two traits sharing the same causal variant (PP_4_) in a 150kb LD window, with PP_4_>0.70 corresponding to strong evidence of colocalisation, as previously suggested [31,32]. Priors chosen for the colocalisation analyses were, p1=10^−3^, p2=10^−4^, and p12=10^−5^, or approximately a 75% prior belief that a signal will only be observed in the LTL GWAS and less than 0.01% prior belief in favor of colocalisation between the two traits at a given locus [33]. Conditioning and masking colocalisation methods were also used as they may identify putative shared causal variants in the presence of multiple causal variants present in a defined LD window [34]. We present the average PP4 from all methods as our posterior belief in favor of colocalisation between LTL and lung cancer risk. Multi-trait colocalisation based on a clustering algorithm was also performed using HyPrColoc (v1.0) to identify shared genetic signals with other lung cancer-related traits [31].

### Principal components analyses based on RNA-sequencing data

Read counts of RNA-sequencing data were normalized within (GC-content and gene length) and between (sequencing depth) lane procedures by EDASeq R package (version 2.28.0) [35] and excluding low read counts. Principal component analysis was applied using singular value decomposition method, after excluding extreme outliers. Pathway analyses were conducted using Gene Set Enrichment Analysis software (GSEA, version 4.2.3) [36] on gene annotations from Gene Ontology database. Pathway analyses were restricted to the top 500 genes positively and negatively correlated with each principal component that passed multiple-testing correction (Bonferroni-adjusted p-value < 0.05 for 74,465 tests), which is the maximum number of genes supported by the software.

The polygenic risk score (PRS) for LTL was comprised of the same 144 variants used in the MR analysis and was computed as the sum of the individual’s beta-weighted genotypes using PRSice-2 software [37]. Associations were estimated per standard deviation increase in the PRS, which was normalized to have a mean of zero across lung adenocarcinoma samples of European ancestry within the TCGA cohort. The associations between the eigenvalues of the gene expression principal components (outcome) and demographic, clinical, and genomic features related to genome stability (predictors derived from TCGA published papers and TCGA data portal, except for the DNA mutational signatures [21]), were calculated using a multivariate linear regression model.

## Results

### Genome-wide genetic correlations

We first assessed the shared genetic basis of telomere length, lung cancer risk, and other putative lung cancer risk factors, such as smoking behaviors (age start smoking, smoking cessation, smoking initiation, and cigarettes per day) and lung function (FEV_1_ and FVC) using genome-wide correlations (Fig. 1A). There was little evidence for genetic correlations by LDSC between LTL variants and lung cancer (r_g_=-0.01, p=0.88) or when stratified by histologic subtypes (Fig. 1A). Increasing LTL was genetically correlated with older age at smoking initiation (r_g_=0.12, p=3.0×10^−3^), and negatively correlated with smoking cessation: (r_g_=-0.21, p=6.9×10^−09^), smoking initiation (r_g_=-0.16, p=1.3×10^−10^), and cigarettes per day (r_g_=-0.19, p=2.1×10^−08^). Longer LTL was genetically correlated with improved lung function, as indicated by increasing values of FEV_1_ (r_g_=0.09, p= 5.1×10^−07^) and FVC (r_g_=0.09, p=1.1×10^−05^). To better understand the absence of genome-wide correlations between LTL and lung cancer, we visualized the Z-scores for each trait for approximately 1.2 million variants included in the LDSC analyses (Fig. 1B). A subgroup of variants associated with longer LTL were correlated with increased lung adenocarcinoma risk, while the subgroup of smoking-behavior associated variants, which also conferred an increased risk of lung adenocarcinoma, tended to have lower LTL.

**Figure 1.**
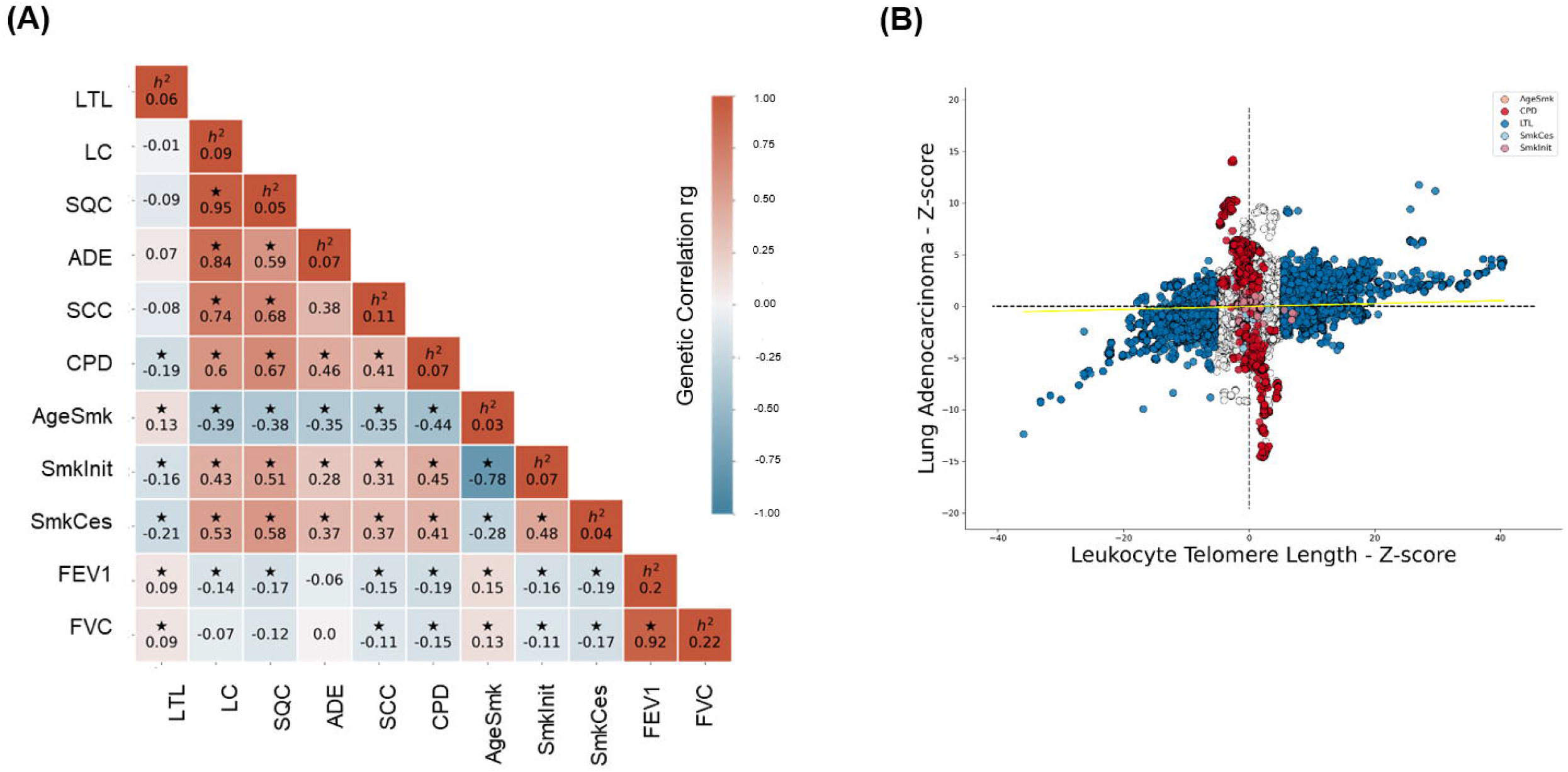
Genetic correlations between leukocyte telomere length (LTL) and lung cancer (LC) related traits. (A) Heatmap representing the genetic correlation analysis (rg) for LTL (first column) across lung cancer (LC), histological subtypes (lung adenocarcinoma (ADE), squamous cell carcinoma (SQC), and small cell carcinoma (SCC)), smoking propensity (cigarettes per day (CPD), Smoking cessation (SmkCes), Smoking initiation (SmkInit), and Age of smoking initiation (AgeSmk)), and lung function related (forced vital capacity (FVC) and forced expiratory volume (FEV1)) traits. The black star indicates correlations that passed Bonferroni correction (p<4e-^04^). Heritability (h^2^) as the proportion of the phenotypic variance caused by SNPs. (B) Plot of Z-scores (ADE vs. LTL), including all the Hapmap (∼1.2 million) SNPs but excluding HLA region. Genome-wide significant SNPs (p<5e-08) for each trait were colored (CPD in red, SmkInit in light red, LTL in blue, SmkCes in lightblue, and not genome-wide hits in white). Linear Regression line was colored in yellow.

### Mendelian randomization analyses

From the 490 genetic instruments associated with LTL at genome-wide significance (p<5e^-08^), 144 LTL genetic instruments, that explained ∼ 3.5% of the variance in LTL, and were in low linkage disequilibrium (r^2^ < 0.01) were used in MR analysis. A polygenic risk score (PRS) comprised of these genetic instruments was associated with TL estimated from whole genome sequencing in blood samples across TCGA cohorts (Beta=0.03, 95%CI=0.01-0.05, p=0.001), but was not associated with TL in tumor material from the same patients (Additional file 1: Fig. S1).

MR analyses demonstrated that longer genetically predicted LTL is associated with increased lung cancer risk (OR=1.62, 95%CI=1.44-1.84, p=9.91×10^−15^) (Fig. 2, Additional file 2: Table S1). Longer LTL conferred the largest increase in risk for lung adenocarcinoma tumors (OR=2.43, 95%CI=2.02-2.92, p=3.76×10^−21^), but there was limited evidence of a causal relationship for other histologic subtypes, such as squamous cell carcinoma (OR=1.00, 95%CI=0.84-1.19, p=0.98) and small cell carcinoma (OR=1.13, 95%CI=0.87-1.45, p=0.34) (Fig. 2, Additional file 2: Table S1). When stratifying the analyses by smoking status, LTL was associated with lung cancer risk in both never (OR=2.02, 95%CI=1.45-2.83, p=3.78×10^−05^) and ever smokers (OR=1.54, 95%CI=1.34-1.76, p=7.75×10^−10^) (Fig. 2, Additional file 2: Table S1).

**Figure 2.**
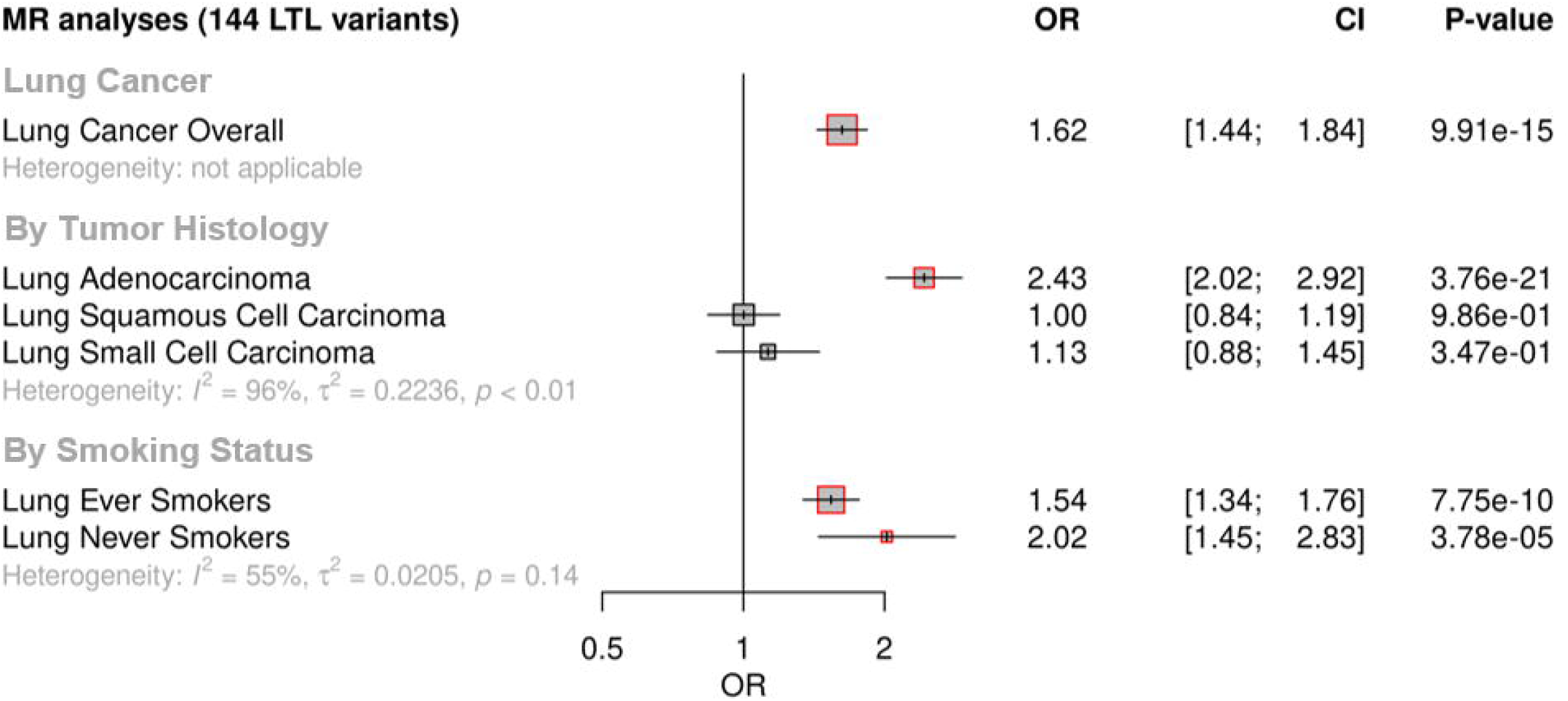
Genetically predicted leukocyte telomere length (LTL) association with lung cancer. Lung cancer (overall, by histology or by smoking status) risk associations with the LTL instrument from the inverse-variance-weighted MR analyses are expressed as odds ratio (OR) per standard deviation increase in genetically predicted LTL. Statistically significant associations with p-values<0.05 (red square). Heterogeneity is estimated by the statistic I^2^, tau variance of subgroups (τ^2^), and p-values for Cochran’s Q heterogeneity measure.

Evidence for negative pleiotropy (Additional file 2: Table S2) and heterogeneity (Additional file 2: Table S3) was observed for all lung cancer outcomes except for squamous cell carcinoma. However, a significant association for LTL and lung cancer risk was found for methods robust to the significant directional pleiotropy (MR-Egger: lung cancer overall [OR=2.35, p=3.13×10^−13^]; lung adenocarcinoma [OR=4.48, p=7.30×10^−17^]; never smokers [OR=6.84, p=2.07×10^−10^]) (Additional file 2: Table S1). Leave-one-out analyses detected only one outlier, rs7705526 in *TERT*, resulting in >10% change in MR effect size for associated lung cancer subtypes (Additional file 2: Table S4). MVMR analyses considering instruments related to LTL and smoking behavior, such as smoking initiation and cigarettes per day, suggested that the association between LTL and lung adenocarcinoma risk is independent of smoking propensity (Additional file 2: Table S5).

### Colocalisation analyses

We investigated whether there was evidence of shared genetic signals between LTL and lung adenocarcinoma at loci centered on the 144 genetic instruments used in MR analyses using colocalisation (Fig. 3A and Additional file 2: Table S6). Loci with evidence of colocalisation between LTL and lung adenocarcinoma tended to be in genes that encode telomerase subunits and its associated complex, including genetic variants at *TERT* (5p15.33) (rs33977403, rs7705526, rs61748181, rs71593392, and rs140648021), *TERC* (3q26.2) (rs12638862 and rs146546514), and *OBFC1* (10q24.33) (rs9419958 and rs139122544). Several colocalised loci mapped to genes that have not been previously linked to lung cancer risk: *MPHOSPH6* (16q23.3) (rs2303262), *PRPF6* (20q13.33) (rs80150989), and *POLI* (18q21.2) (rs2276182).

**Figure 3.**
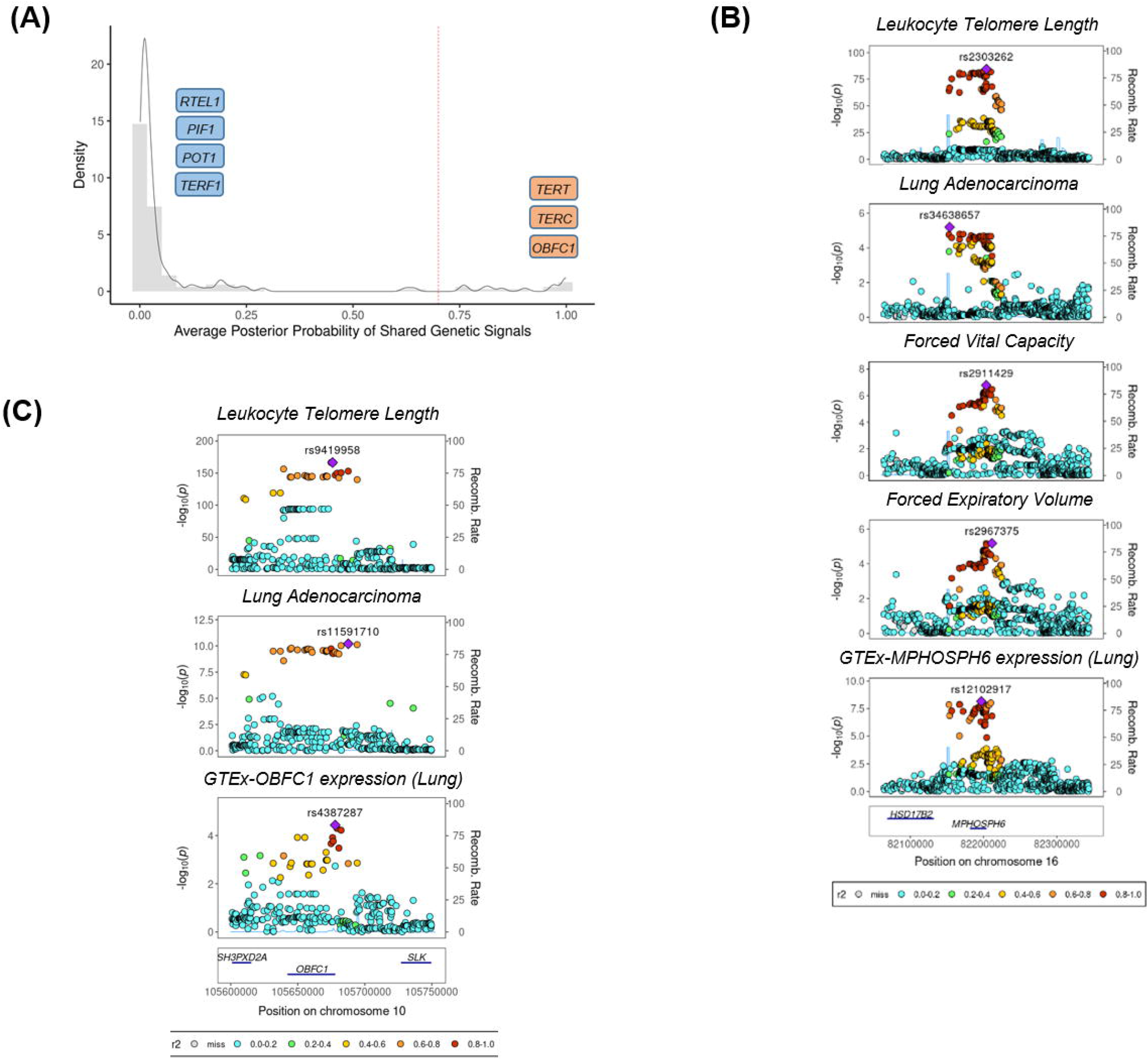
Colocalisation analyses for the genetic loci defined by the 144 LTL variants. (A) Distribution of the average posterior probability for shared genetic loci between leukocyte telomere length and lung adenocarcinoma, highlighting in orange the telomere maintenance loci that colocalised (avg_PP4 ≥0.70) and in blue the ones where there was limited evidence for colocalisation (avg_PP4<0.70). Dashed red line represent the arbitrary avg_PP4 cutoff of 0.70. Representative stack plots for the multi-trait colocalisation results within (B) *MPHOSPH6* and (C) *OBFC1* loci, centered on a 150kb LD window of rs2303262 and rs9419958 variants, respectively. Left Y-axis represents the –log10(p-values) of the association in the respective genome-wide association study for a given trait. The right Y-axis represents the recombination rate for the genetic loci. The X-axis represents the chromosome position. SNPs are colored by the linkage disequilibrium correlation threshold (r2) with the query labeled SNP in European population. Sentinel SNPs within the defined LD window were labelled in each trait.

Other telomere maintenance genes showed limited evidence of colocalisation with lung adenocarcinoma (i.e, *TERF1* and *PIF1*). For instance, while the *RTEL1* locus (20q13.33: rs117238689, rs115610405, rs35640778, and rs35902944) harbored variants associated with both LTL and lung adenocarcinoma (Additional file 1: Fig. S2 and Additional file 2: Table S6), these signals appeared to be distinct and independent of each other (Fig. 3A and Additional file 2: Table S6).

We further evaluated whether the loci colocalised between LTL and lung adenocarcinoma also shared genetic signals with other traits related to lung cancer susceptibility (Additional file 2: Table S7). Multi-trait analyses at the 16q23.3 locus colocalised rs2303262 with *MPHOSPH6* expression in lung tissue, FVC and FEV_1_, but not with any of the traits related to smoking behavior (PP=0.72) (Fig. 3B and Additional file 2: Table S7). We additionally identified evidence of colocalisation (PP=0.74) between lung adenocarcinoma, LTL, and gene expression in lung epithelial cells for two variants at the *OBFC1* locus: rs139122544 and rs9419958 (Fig. 3C and Additional file 2: Table S7).

### Genetically predicted LTL association with tumor features

We investigated the impact of genetically predicted LTL on lung adenocarcinoma tumor features by estimating molecular expression patterns within 343 lung adenocarcinomas tumors using principal component analysis in RNA-sequencing data. The first 5 components explained ∼54% of the observed variance in the RNA-sequencing data (Fig. 4 and Additional file 1: Fig. S3A,B). To explore the biological meaning of the five components, we performed pathway analyses for the top 500 genes with the highest loadings in each component (Additional file 2: Tables S8-S9). Overall, the genes correlated with each component tended to be enriched for specific cell signaling pathways (PC1: RNA processing; PC2: Cell-cycle; PC3: Metabolic processes; PC4: Immune response; PC5: Cellular response to stress and DNA damage; False Discovery Rate < 5%) (Additional file 2: Table S10).

**Figure 4.**
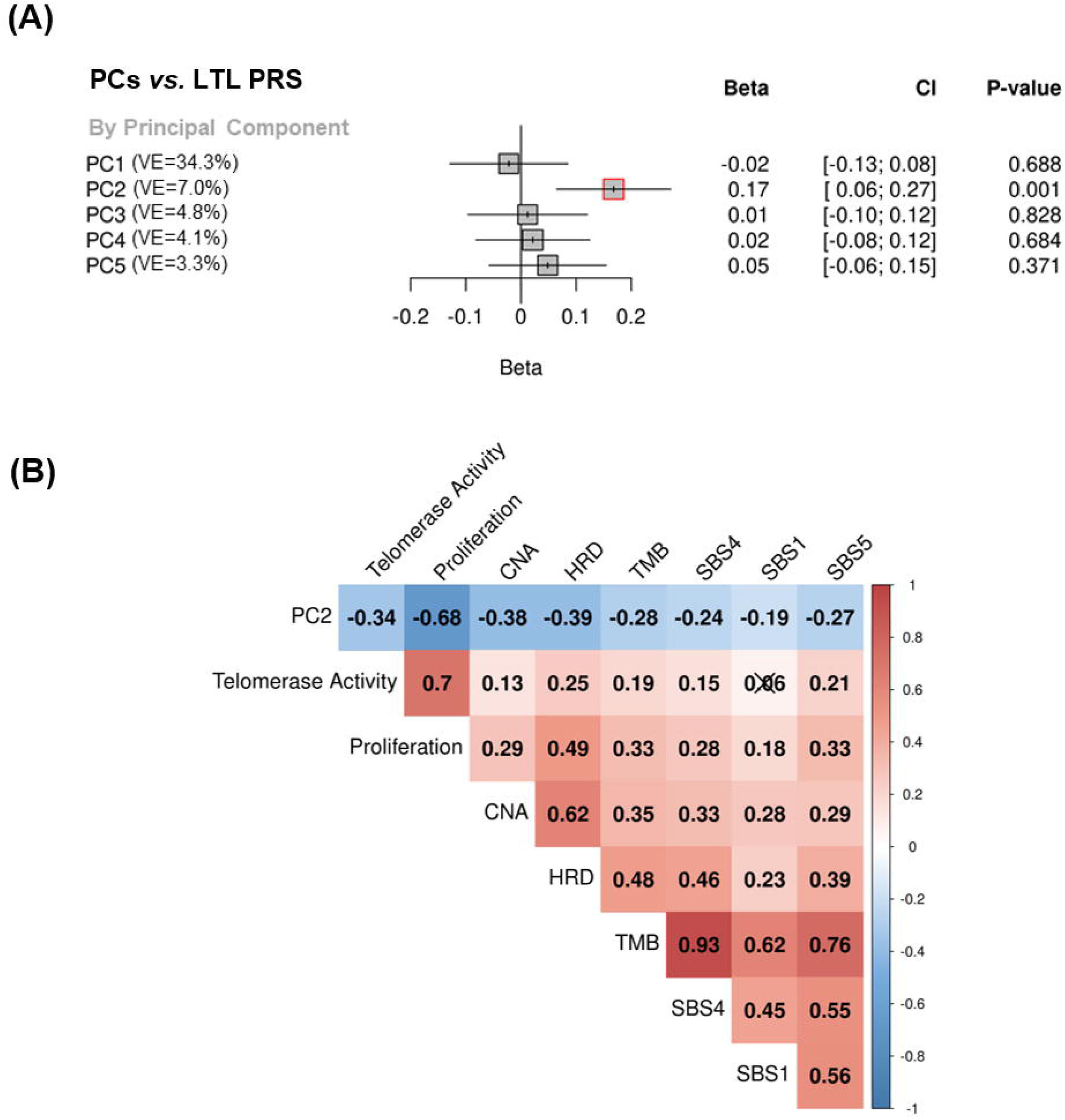
Associations between molecular expression patterns of lung adenocarcinoma tumors, LTL PRS, and TCGA features. (A) LTL PRS association with the first 5 principal components based on RNA-sequencing data of lung adenocarcinomas tumors. Results are expressed as beta estimate per standard deviation increase in genetically predicted LTL. Linear regression model adjusted by sex, age, smoking status, and PC1-5 (genetic ancestry) covariates. Statistically significant associations with p-values<0.05 (red square). VE= variance explained of eigenvector within RNA-sequencing data from lung adenocarcinomas (B) Heatmap representing the correlations among PC2 and selected molecular features related to telomere length canonical roles. LTL=leukocyte telomere length; PRS=polygenic risk score; PC=principal component; TMB=tumor total mutation burden; HRD=Homologous Recombination Deficiency, SBS (single base substitution DNA mutational signatures). SBS1 and SBS5 are DNA mutational signatures associated with age-related processes and SBS4 is associated with tobacco smoking exposure. X-shaped marker to cross correlations with p-value>0.05.

We then tested the association between the polygenic risk score comprised of the 144 genetic instruments selected for MR analysis and the five components of gene expression within lung adenocarcinoma tumors (Fig. 4A). The LTL PRS was positively associated with the second component (PC2) of tumor expression (Beta=0.17, 95%CI=0.12-0.19, p=1.0×10^−3^) (Fig. 4A). In multivariate analysis, higher values of PC2 tended to be associated with patients older at diagnosis (p=0.001), female (p=0.005), being never smokers (p=0.04), and diagnosed with early-stage tumors (p=0.002) (Table 1). PC2 was also highly correlated with gene expression-based measure of cell proliferation and several genomic features related to genomic stability (Fig. 4B). In multivariate analysis, higher values of PC2 were associated with reduced tumor proliferation (p=3.7×10^−30^), lower somatic copy number alternations (p=1.6×10^−05^), and higher tumor telomerase activity scores (p=1.6×10^−5^). Multivariate analysis also indicted that LTL PRS remained an independent predictor of PC2 when considering these genomic features (p=0.009) (Table 1). It is noteworthy only nominal associations between LTL PRS and above-mentioned features, and none remained statistically significant after correction for multiple testing (Additional file 2: Table S11).

**Table 1.**
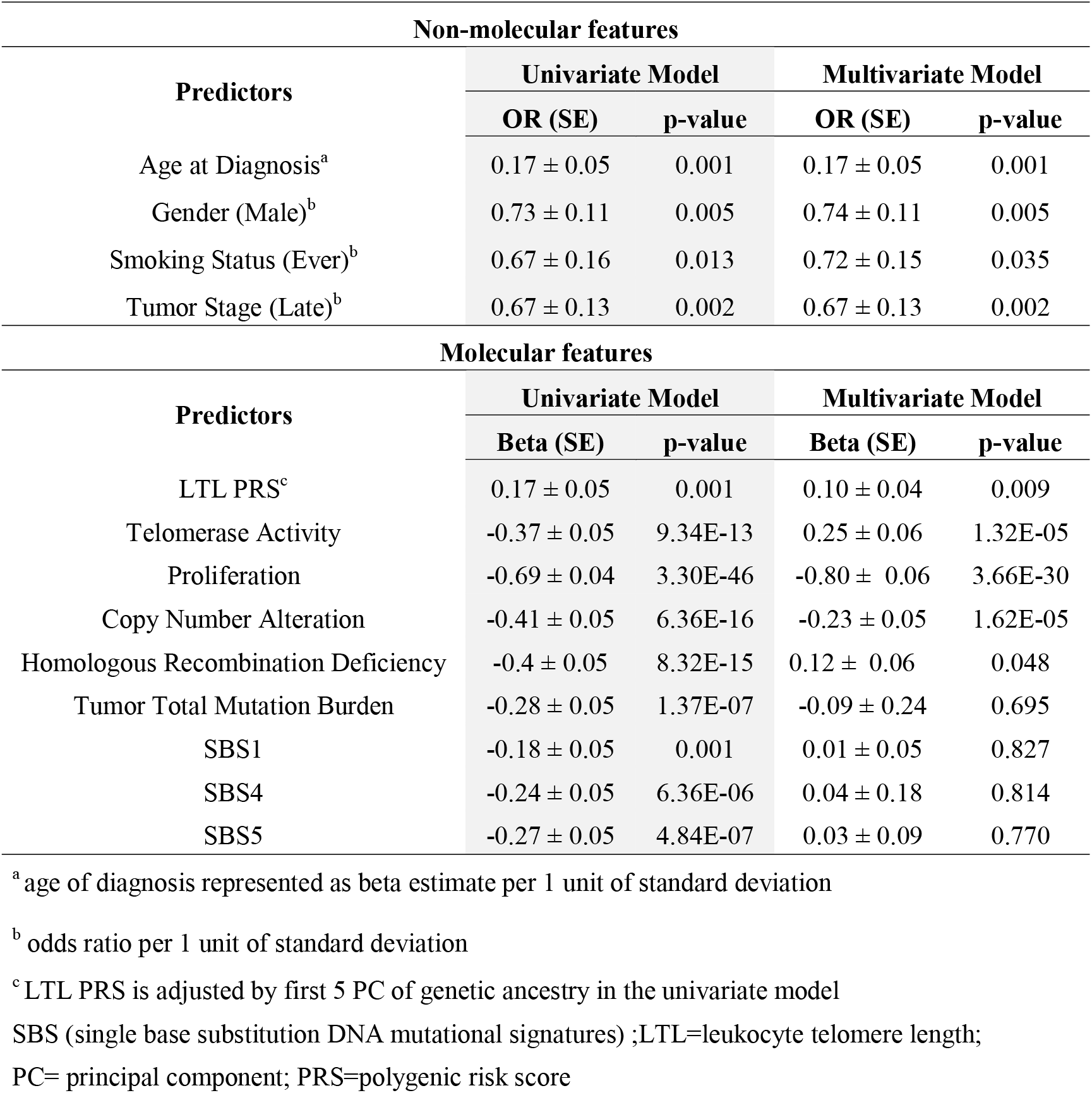
Association between PC2 (outcome) and lung adenocarcinoma tumor features in univariate and multivariate models (n=343).

| Non-molecular features |  |  |  |  |
| --- | --- | --- | --- | --- |
| Predictors | Univariate Model |  | Multivariate Model |  |
|  | OR (SE) | p-value | OR (SE) | p-value |
| Age at Diagnosis<sup>a</sup> | 0.17 ± 0.05 | 0.001 | 0.17 ± 0.05 | 0.001 |
| Gender (Male)<sup>b</sup> | 0.73 ± 0.11 | 0.005 | 0.74 ± 0.11 | 0.005 |
| Smoking Status (Ever)<sup>b</sup> | 0.67 ± 0.16 | 0.013 | 0.72 ± 0.15 | 0.035 |
| Tumor Stage (Late)<sup>b</sup> | 0.67 ± 0.13 | 0.002 | 0.67 ± 0.13 | 0.002 |
| Molecular features |  |  |  |  |
| Predictors | Univariate Model |  | Multivariate Model |  |
|  | Beta (SE) | p-value | Beta (SE) | p-value |
| LTL PRS<sup>c</sup> | 0.17 ± 0.05 | 0.001 | 0.10 ± 0.04 | 0.009 |
| Telomerase Activity | -0.37 ± 0.05 | 9.34E-13 | 0.25 ± 0.06 | 1.32E-05 |
| Proliferation | -0.69 ± 0.04 | 3.30E-46 | -0.80 ± 0.06 | 3.66E-30 |
| Copy Number Alteration | -0.41 ± 0.05 | 6.36E-16 | -0.23 ± 0.05 | 1.62E-05 |
| Homologous Recombination Deficiency | -0.4 ± 0.05 | 8.32E-15 | 0.12 ± 0.06 | 0.048 |
| Tumor Total Mutation Burden | -0.28 ± 0.05 | 1.37E-07 | -0.09 ± 0.24 | 0.695 |
| SBS1 | -0.18 ± 0.05 | 0.001 | 0.01 ± 0.05 | 0.827 |
| SBS4 | -0.24 ± 0.05 | 6.36E-06 | 0.04 ± 0.18 | 0.814 |
| SBS5 | -0.27 ± 0.05 | 4.84E-07 | 0.03 ± 0.09 | 0.770 |

<sup>a</sup> age of diagnosis represented as beta estimate per 1 unit of standard deviation
<sup>b</sup> odds ratio per 1 unit of standard deviation
<sup>c</sup> LTL PRS is adjusted by first 5 PC of genetic ancestry in the univariate model
SBS (single base substitution DNA mutational signatures) ;LTL=leukocyte telomere length;
PC= principal component; PRS=polygenic risk score

## Discussion

The maintenance of telomere length is one of the hallmarks of cancer, being critical for cell proliferation and genome integrity [4]. Individual differences in telomere length, measured either directly or indirectly by germline determinates has been linked with multiple diseases, including cancer susceptibility [7]. The measurement of LTL within the UKBB has provided as resource for the development of a more powerful set of genetic instruments that capture a greater proportion of variation in LTL compared to previous studies [7]. We applied genetic determinants of LTL to the largest GWAS of lung cancer to further characterize the role of telomere maintenance in lung cancer etiology.

Using an MR analysis framework, we confirmed the previously reported relationship between genetically predicted longer LTL and increased risk of lung cancer. Our expanded genetic instrument detected systematic negative pleiotropy, which has not been observed in previous MR studies [14,15,16]. Correcting for this pervasive directional bias resulted in substantially larger effects of LTL on risk of lung adenocarcinoma and lung cancer in never smokers, implying that LTL may even be more important to these phenotypes than previously estimated [14,15,16]. Our observations in never smokers was also supported by multivariate MR analyses where adjustment for smoking did not attenuate the effect of LTL on lung cancer susceptibility.

Colocalisation analyses within the variants selected with the MR genetic instrument highlighted shared genetic signals between LTL and lung adenocarcinoma, including loci that contain telomere maintenance related genes (*TERT, TERC*, and *OBFC1*) and three genetic loci not previously linked with lung cancer susceptibility (*POLI, PRPF6*, and *MPHOSPH6*). The lung cancer risk allele of the *MPHOSPH6* sentinel variant (rs2303262) was associated with longer LTL, reduced pulmonary function, and increased *MPHOSPH6* gene expression in lung tissue. *MPHOSPH6* encodes an enzyme associated with the RNA exosome complex where it modulates RNA binding activity. *PRPF6* in 20q13.33 is involved in androgen binding and has been shown to promote colon tumor growth via preferential splicing of genes involved in proliferation [38]. *POLI* is a member of the Y-family of DNA damage-tolerant polymerases involved in translesion synthesis [39]. As part of its role in DNA repair and replication stress, *POLI* interacts with *TP53* to bypass barriers during DNA replication, which may confer a pro-survival effect to stem cells and cancer cells [40]. Colocalisation also highlighted the heterogeneity in the genetic effects of LTL loci, with several loci important for telomere maintenance not colocalised with lung cancer.

We additionally identified the relationship between genetic determinants of LTL and a specific gene expression component in lung adenocarcinoma tumors. The aspect of this component associated with longer LTL, which we note above is associated with increased lung cancer risk, was also associated with demographic and clinical features, such as never smoking, female and early-stage tumors compared with other lung adenocarcinoma patients. This expression component also tended to be related to genomic features related to genomic stable tumors and strikingly associated with cell proliferation score, implying that this component might be a proxy for this feature. These results appear consistent with the canonical role of telomere length in preserving genome stability and cell proliferation [1]. The fact that long genetically LTL was associated with increased risk of lung cancer in our MR analyses could also be reasonably explained by the enhanced clonal expansion and higher probability of accumulating driver events in individuals with longer LTL at the initial steps of carcinogenesis, as previously observed in other cancers [41].

Despite the robust and large effects of LTL on lung cancer risk observed in MR, the genetic correlation between LTL and lung cancer was effectively null. The LDSC approach considers genetic variants across the entire genome, whereas the MR approach preferentially selects variants based on their association with LTL, restricting to those that achieved genome-wide significance. One possibility for the lack of genetic correlation between LTL and lung cancer is that genetic variants may differ in the direction that they influence these traits. For example, the subgroup of genetic variants noted at genome-wide significance from LTL studies was associated with increased LTL and LC risk. However, the subgroup related to smoking behaviors which, in turn, are linked with increased LC risk, tends to decrease LTL. If such opposing effects were widespread across the genome, it could account for the lack of genetic correlation between LTL and lung cancer estimated by LDSC and highlights the complex nature of the genetic variants that determine LTL and lung cancer risk.

Some limitations of this study should be acknowledged. Our colocalisation approach is generally more conservative and may fail to accurately determine the posterior probability for shared genetic signals in the presence of multiple independent associations in a given locus [42], which may be a plausible explanation for the lack of colocalisation observed at *RTEL1* locus. Furthermore, the relatively small sample size of the lung adenocarcinoma cohort from TCGA may have reduced the power of our study, and larger cohorts of expression profiles tumors will be necessary to validate and explore some of our findings. The potential limitations such as collider bias within the lung adenocarcinoma case only study design should also be considered.

## Conclusions

In conclusion, we describe an association between long genetically predicted LTL and lung cancer risk, which provides insights into how telomere length influences the genetic basis of lung cancer etiology. By using a novel framework to explore the biological implications of genetically complex traits, we unravel one gene expression component, highly correlated with proliferation rate score and other genomic stability-related features, associated with LTL in lung adenocarcinoma tumors. These findings suggest that lung adenocarcinoma patients with longer LTL might have more genomic stable tumors than the ones with shorter LTL, shedding some light on telomere biology in those tumors.

## Data Availability

Lung cancer summary statistics obtained from ILCCO can be accessed by the database of Genotypes and Phenotypes (dbGAP) under accession phs000876.v1.p1. The GWAS summary statistics for tobacco-smoking behaviors (GSCAN: https://conservancy.umn.edu/handle/11299/201564), LTL (https://figshare.com/s/caa99dc0f76d62990195), and GTEx version 8 (downloaded via GTEx google cloud resource) are publicly available. TCGA data was accessed by dbGAP through Project #2731. RNA-sequencing data and TCGA-related data are publicly available as described in the data section. The data generated in this study are available within the article and its supplementary data files.

## Abbreviations

FEV_1_: Forced expiratory volume in 1 second
FVC: Forced vital capacity
GWAS: Genome-wide association studies
LTL: Leukocyte telomere length
LDSC: Linkage disequilibrium score regression
MR: Mendelian randomization
PCA: Principal component analysis
PP: Posterior probability
PRS: Polygenic risk score
UKBB: UK Biobank
TCGA: The Cancer Genome Atlas

## Declarations

### Ethics approval and consent to participate

Not applicable

### Consent for publication

Not applicable

## Authors’ contributions

The project was conceived and supervised by J.D.M. R.C.C.P. participated in the Mendelian Randomization analyses, conducted the colocalisation and principal component analyses, wrote and reviewed the manuscript. K.S. and L.K. performed the Mendelian Randomization analyses and participated in the drafting, writing, and reviewing of the manuscript. J.R.A. performed the LD score regression analyses and participated in the drafting and reviewing of the manuscript. V.C., N.J.S., C.P.N. provided the LTL GWAS summary statistics and participated in the review of the manuscript. M.M. and A.A.G.G. performed the quality control of the germline variants of the TCGA dataset and participated in the reviewing of the manuscript. P.H, S.K, C.A, P.B, R.J.H participated in the reviewing of the manuscript. All authors read and approved the final manuscript.

## Acknowledgements

We would like to acknowledge the TCGA Research Network (https://www.cancer.gov/tcga) and the contribution of specimen donors and research groups involved in this resource. We also would like to acknowledge the ILLCO consortium, the participants of the UK biobank and GTEx project and the supporting bodies (https://commonfund.nih.gov/GTEx), specimen donors and research groups.

## Disclaimer

Where authors are identified as personnel of the International Agency for Research on Cancer/World Health Organization, the authors alone are responsible for the views expressed in this article and they do not necessarily represent the decisions, policy, or views of the International Agency for Research on Cancer/World Health Organization.

## Supplementary Information

***Additional File 1: Figures S1-S3***

**Figure S1**. Association between genetically predicted leukocyte telomere length (LTL) with measured telomere length in TCGA cohorts. **Figure S2**. Association plots for LTL and lung adenocarcinoma at RTEL1 locus. **Figure S3**. PCA analyses based on RNA-sequencing data.

***Additional File 2: Tables S1-S11***

**Table S1**. Mendelian randomization analyses across methods for the 144 leukocyte telomere length SNPs. **Table S2**. Test for directional pleiotropy using MR Egger for the 144 leukocyte telomere length SNPs. **Table S3**. Heterogeneity tests for the 144 leukocyte telomere length SNPs. **Table S4**. Leave-one out analyses for the 144 leukocyte telomere length SNPs. **Table S5**. Multivariable Mendelian randomization analyses. **Table S6**. Summary of the colocalisation results for the LTL instrument based on 144 variants. **Table S7**. Summary of the Multi-trait colocalisation results for the 12 colocalised genetic loci. **Table S8**. Top 500 genes positively correlated with the first 5 principal components. **Table S9**. Top 500 genes negatively correlated with the first 5 principal components. **Table S10**. Pathway Analysis on the top 500 genes positively and negatively correlated with the first 5 principal components based on RNA-seq data of 343 ADE cases from TCGA datase using GSEA. **Table S11**. Association of LTL PRS with the clinical and molecular features of lung adenocarcinoma tumors.

