## Supplementary Figures S1-S3 for "Common genetic variations in telomere length genes and lung cancer"

**Supplementary Materials**

**Supplementary Figures**

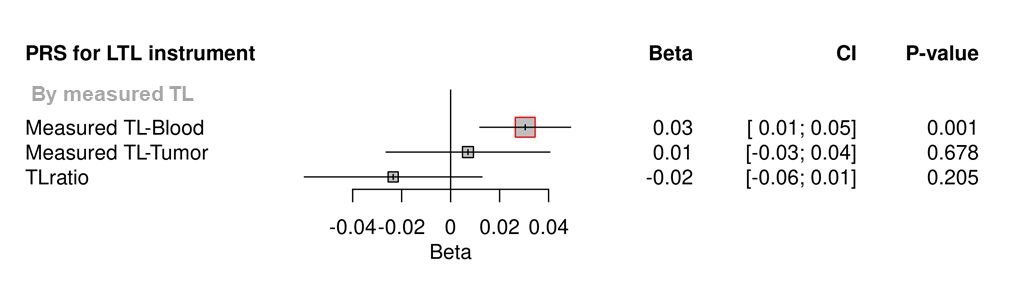

**Figure S1.** **Association between genetically predicted leukocyte telomere length (LTL) with measured telomere length in TCGA cohorts.** Telomere length (TL) was measured by Barthel et al. in a subset of high-confident samples from TCGA cohorts using whole-genome sequencing (n=655). TL was directly measured in blood and tumor samples, and log(tTL/nTL) (TLratio) were also obtained, from several TCGA cohorts. Associations were expressed as beta estimate per standard deviation longer LTL in log scale. P-values<0.05 (red square). Sex, age at diagnosis, cohort, and principal components (PC1-5) were used as covariates in the linear regression model.

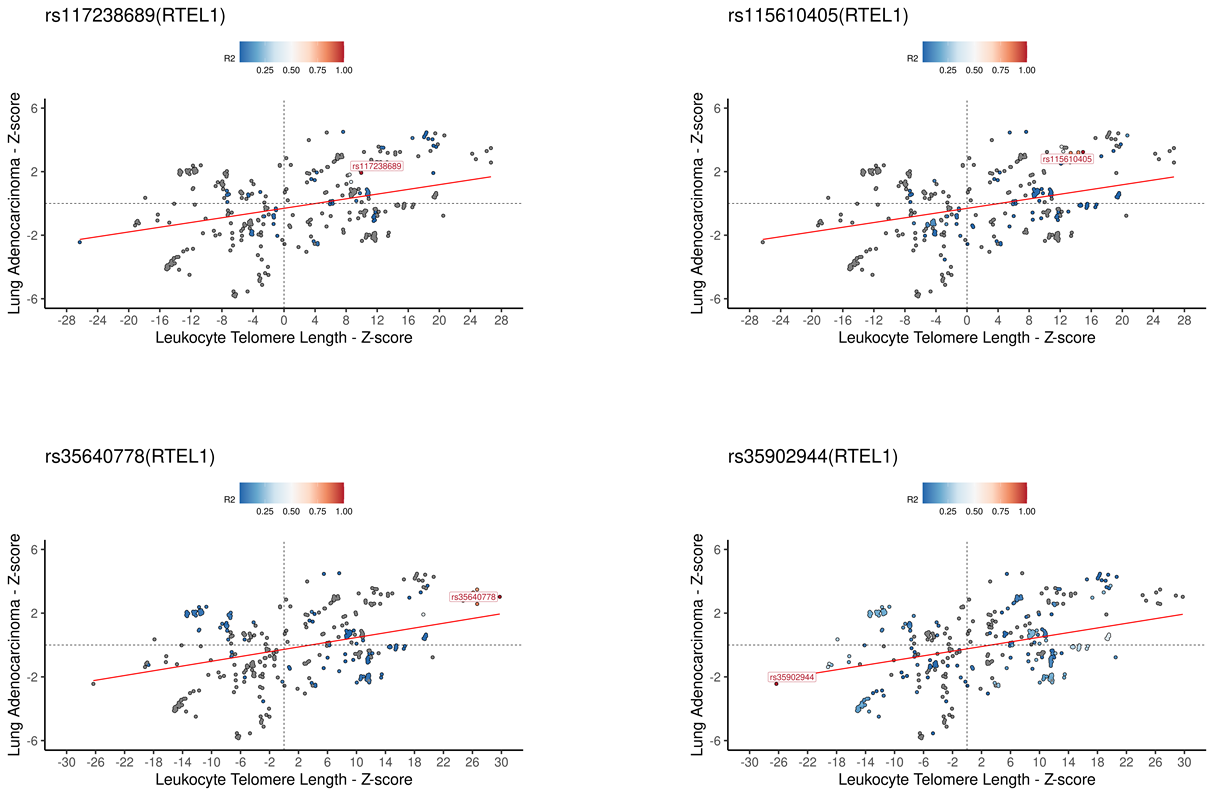

**Figure S2. Association plots for LTL and lung adenocarcinoma at *RTEL1* locus.** Z-score plots for genetically predicted LTL and lung adenocarcinoma risk for the 4 LTL variants annotated for *RTEL1*. The genetic variants were colored by the linkage disequilibrium correlation threshold (r^2^) with the query labeled SNP in a defined LD window of 150kb centered on the query SNP in European population. Z-score defined as the beta estimate divided by standard error for each SNPs in the respective genome-wide association study.

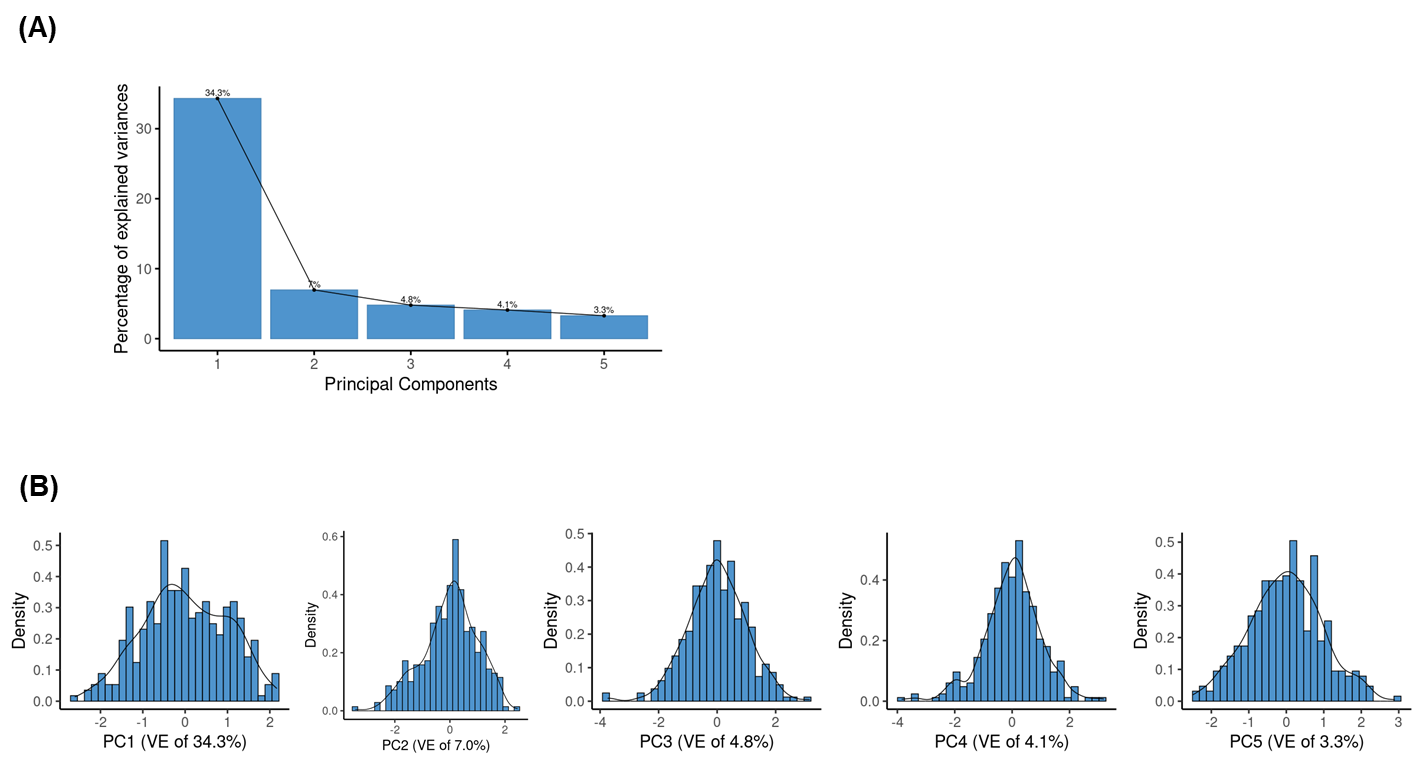

**Figure S3. PCA analyses based on RNA-sequencing data.** The RNA sequencing data from 343 primary lung adenocarcinoma tumor samples were retrieved. (A) Principal components analysis was applied to the centered log-transformed gene read counts, and the first 5 principal components were represented, which explained 53.5 % of the variance in the gene expression for those samples. (B) The distributions of the first 5 principal components are represented in the density plots.
